# Omega-3 & Aspirin Boost in Periodontal Therapy for Type II Diabetes: A Systematic Review

**DOI:** 10.1101/2024.10.10.24315256

**Authors:** Emma Walker, John Franklin, Jonathan Robinson

## Abstract

**Aim:** Omega-3 polyunsaturated fatty acids and aspirin are increasingly explored as adjuncts in periodontal therapy for patients with type II diabetes, owing to their anti-inflammatory properties. Despite preliminary evidence suggesting their effectiveness in reducing periodontal inflammation and improving metabolic control, comprehensive evaluations at the clinical level remain limited. This systematic review seeks to critically assess the existing evidence to determine the efficacy of these agents in modifying clinical, biochemical, and metabolic parameters in this high-risk patient group.

**Methods:** A comprehensive search was conducted across electronic databases, including CINAHL, MEDLINE, PubMed, Embase, Dentistry & Oral Sciences Source, and Cochrane Library, to identify relevant studies from inception to January 22, 2024. Included studies were required to be randomised controlled trials and comprise both omega-3 and aspirin as an adjunct to non-surgical treatment. Data extraction was completed independently, and accuracy confirmed by a second reviewer. Data were synthesised using the revised Cochrane risk of bias tool for randomised trials.

**Results:** A total of 76 records were identified, and three studies met the inclusion criteria. One demonstrated a statistically significant difference in pocket depth, clinical attachment gain and inflammatory markers compared to the control group, and two showed a significant improvement in glycated haemoglobin levels.

**Conclusion:** Omega-3 and aspirin combined have the potential to improve clinical parameters and reduce inflammatory markers in the gingival crevicular fluid. Following non-surgical periodontal treatment, this combination may enhance glycemic control and aid in managing type II diabetes. This systematic review offers insights into the potential effectiveness of omega-3 polyunsaturated fatty acids and aspirin as adjuncts to non-surgical periodontal therapy in patients with type II diabetes and periodontitis. However, current research is limited by small sample sizes and varied study designs. Consequently, there is insufficient evidence to integrate this approach into clinical practice at present.

## 1. INTRODUCTION

Periodontitis is an inflammatory disease that can lead to progressive destruction of the surrounding periodontal tissues, leading to potential tooth loss.^1^ Plaque formation is needed to initiate periodontal inflammation; however, it is not sufficient to induce periodontal destruction by itself.^2^ Destruction is caused by an exaggerated immune-inflammatory host response to the microbial challenge.^2^ Periodontal pathogens in plaque trigger a pro-inflammatory response involving the recruitment of neutrophils which leads to a local release of reactive free radicals and enzymes which damage the host tissue.^3^ If there is a constant state of imbalance between the host’s immune-inflammatory response and excess bacterial plaque, then disease will occur.^3^

Severe periodontitis is the sixth most prevalent disease worldwide affecting nearly 750 million people.^1^ The Adult Dental Health Survey for England, Wales, and Northern Ireland identified 8% of adults have at least one pocket of >6mm.^4^ Those experiencing severe periodontitis may experience periodontal inflammation, loss of periodontal attachment, bone loss and tooth mobility which can have a negative impact on daily life.^5^

Type II Diabetes Mellitus is a common metabolic disease characterised by a disorder in insulin secretion, insulin resistance or a combination of both resulting in hyperglycaemia.^6^ Prolonged hyperglycaemia causes serious damage to multiple body systems causing disease.^5^ The World Health Organisation estimated there were 422 million adults worldwide with Type II Diabetes Mellitus.^7^ Chee et al.^8^ suggests a bi-directional relationship exists between diabetes and periodontitis. Diabetes does not cause periodontitis but increases the risk for periodontal diseases and in turn may exacerbate worsening glycaemic control.^3^

Glycaemic control is measured by assessing the level of glycated haemoglobin (HbA1c) in the blood, as hyperglycaemia causes glucose to irreversibly bind to the haemoglobin molecule.^5^ HbA1c can be expressed as a percentage which indicates the amount of glucose molecules absorbed onto haemoglobin.^5^ Alternatively, HbA1c can be reported in mmols/mol. Non-diabetics have HbA1c levels around 5.5% (37mmol/mol).^5^ Type II Diabetes Mellitus patients with HbA1C levels of <7.0% (53mmol/mol) show good glycaemic control.^5^ Levels above >8-9% (64-75mmol/mol) indicate poor glycaemic control. Complications of diabetes are linked to the level of glycaemic control; it has been shown that the risk of diabetic complication reduces with each 1% reduction in HbA1c.^5^

Prolonged hyperglycaemia can cause excess glucose in the gingival crevicular fluid (GCF).^8^ Bacteria thrive on sugar and enriched glucose-GCF may alter the bacteria composition within the biofilm which can favour and influence the development of periodontitis.^9^ Diabetics have an impaired neutrophil function, defective chemotaxis and phagocytosis which enables bacterial persistence in the periodontal pocket increasing destruction.^3^ Often patients have a hyper-responsive monocyte-macrophage phenotype which can increase the production of pro-inflammatory cytokines and mediators.^8^ Elevated levels of pro-inflammatory cytokines accumulating in the GCF is related to glycaemic control.^8^ Mealey et al.^6^ demonstrated Type II Diabetes Mellitus periodontal patients with HbA1c levels >8% had double the amount of interleukin-1b (IL-1b) in the GCF compared to Type II Diabetes Mellitus patients with HbA1c levels <8%. An increase in other inflammatory mediators such as tumour necrosis factor-α (TNF-α) result in increased periodontal inflammation, clinical attachment loss and bone loss.^6^

To our knowledge there have been no systematic reviews which have synthesised the findings of studies investigating the use of omega-3 and aspirin in people with Type II Diabetes Mellitus and periodontal disease. It is therefore the aim of this review to evaluate the effectiveness of omega-3 and aspirin as an adjunct to NSPT in people with Type II Diabetes Mellitus and periodontitis in decreasing clinical, biochemical, and metabolic parameters.

## 2. MATERIAL AND METHODS

This Systematic review followed the Preferred Reporting Items of Systematic Reviews and Meta Analyses (PRISMA) guidelines.^10^ The review protocol was registered with PROSPERO, registration number: CRD42022341870 (22 July 2022) at: https://www.crd.york.ac.uk/prospero/display_record.php?ID=CRD42022341870

### 2.1 Literature search strategy

Table 1 presents the items using the PICO (participants/patient, interventions, comparators, outcomes) format.

**Table 1:**
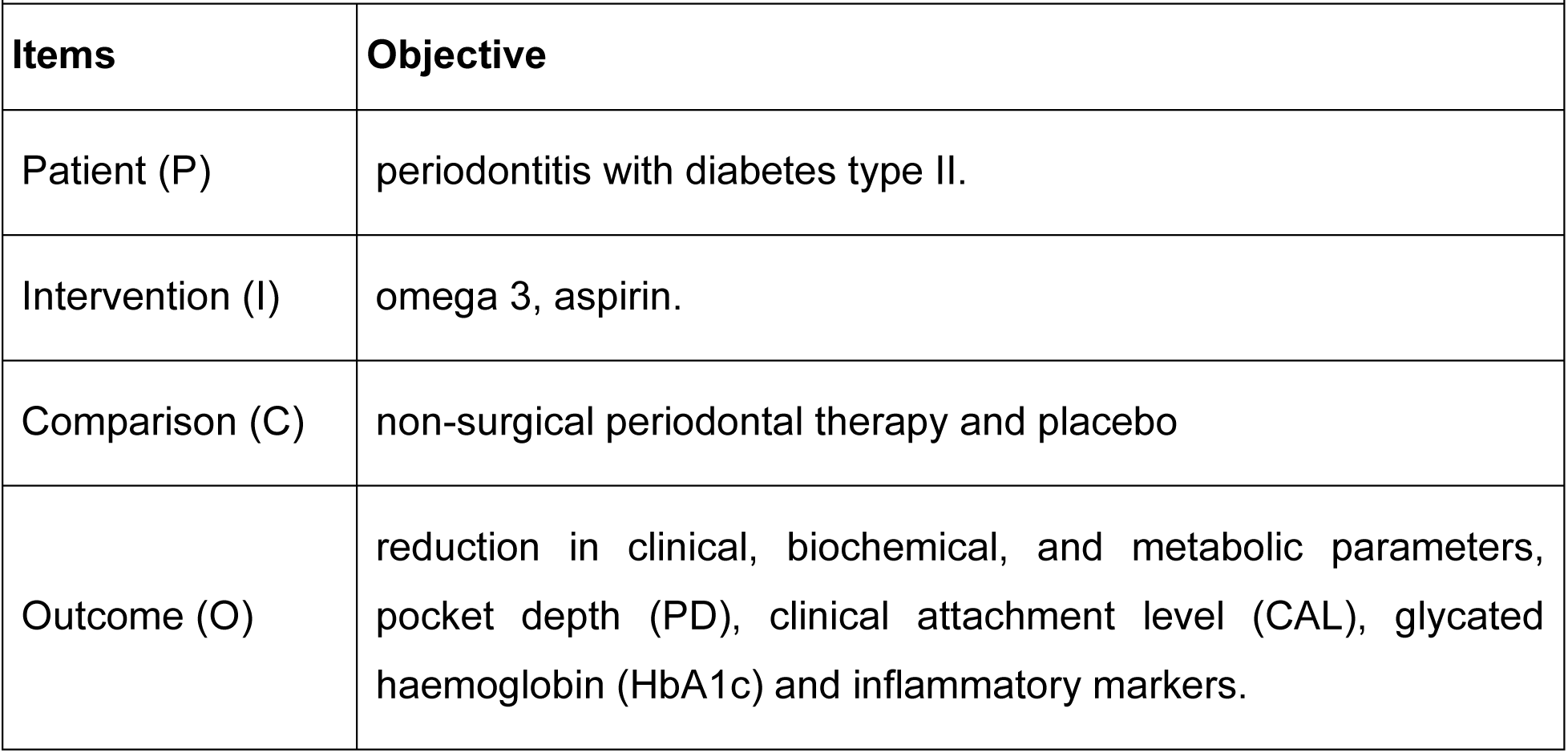
Summary of PICO components. PICO objectives.

### 2.2 Criteria for Selecting Studies into the Review

To be included in this review studies were required to meet the following criteria (summarised in table 2): *Population:* Adults (18 years+) with a confirmed diagnosis of both diabetes mellitus type 2 and periodontal disease; *Intervention:* assess the combination of omega-3 and aspirin; *Comparison:* non-surgical periodontal therapy (NSPT) and placebo groups; *Outcome:* assess clinical, biochemical, and metabolic parameters, pocket depth (PD), clinical attachment level (CAL), glycated haemoglobin (HbA1c) and inflammatory markers; *Types of Studies:* be randomised controlled trials.

**Table 2:** Summary of inclusion and exclusion criteria.

|  | Inclusion criteria | Exclusion criteria |
| --- | --- | --- |
| Population | Adults<br>Type II diabetes<br>Periodontal disease | Gingivitis |
| Intervention | Omega-3 and aspirin | N/A |
| Comparison | non-surgical periodontal therapy<br>placebo | N/A |
| Outcome | Pocket depth<br>Clinical attachment level<br>Glycated haemoglobin (HbA1c) | N/A |
| Types of studies | Randomised controlled trial | Grey literature |

### 2.3 Search strategy

An initial search was conducted by two reviewers (EW, JH) for published systematic reviews followed by a comprehensive search of MEDLINE and PubMed to identify relevant terms and citations. An extensive search was then conducted using MEDLINE, PubMed, Embase, Dentistry & Oral Sciences Source, and CINAHL databases in August 2022. Box 1 shows an example of a MEDLINE search which was run in each database. Box 1 provides an example of a MEDLINE search that was conducted in each database. Hand searching was completed when a relevant study was found in the text and references of the studies that were identified in the database search. On 22^nd^ January 2024, all electronic searches were updated.

#### Box 1

##### Search terms and Boolean operators used in comprehensive search strategy.

[(periodontitis OR periodontal disease) AND (diabetes mellitus OR diabetic type 2 OR diabetic type II) AND (non-surgical periodontal therapy OR scaling OR debridement) AND (omega-3 poly unsaturated fatty acids OR omega 3 OR docosahexanenoic OR eicosapentaenoic) AND (aspirin OR asa OR acetylsalicylic)]

### 2.4 Selection of studies

One reviewer (EW) transferred the found studies to an Excel spreadsheet whereby all duplicates were detected, deleted, and checked manually for accuracy. This was then imported into Rayyan - a web and mobile app for systematic reviews (https://www.rayyan.ai/, ^11^). Two reviewers (EW and JR) screened all abstracts, and then full-text articles, independently and blindly using Rayyan, based on the PICO framework and inclusion and exclusion criteria (see Table 2). Disagreements between the reviewers, would have been discussed with a third reviewer - no disagreements were found. Cohen’s kappa statistic was calculated at each phase of the selection process (title and abstract, and full text) to evaluate agreement between raters.^12^

### 2.5 Data extraction and assessment of risk of bias

The full-text of the selected articles were read and data extracted by one author (EW). Extracted data included: characteristics of the study and population (i.e., age, sex); definition of diabetes (i.e., type, glycaemic control); definition of periodontitis (i.e., moderate, severe); Intervention (i.e., type, duration) and control group (i.e., placebo contents); and follow up time period and measured outcome parameters. Outcome parameters were pocket depth (PD), clinical attachment level (CAL), glycated haemoglobin (HbA1c), interleukin (IL-1β) monocyte chemo-attractive protein (MCP) and pentraxin (PTX3). Included studies were required to have one or more test groups and follow up periods. Where data were reported in a box and whisker plot, the Digitizelt software programme^13^ was used to extract data and converted into median interquartile range. The included studies were assessed by one reviewer (EW) using a standard table based on the revised Cochrane risk of bias 2.0 tool for randomised trials (RoB 2; ^14^).

### 2.6 Data Synthesis

The synthesis without Meta-Analysis (SWiM) guidelines were used to underpin the synthesis approach.^15^ A narrative synthesis approach was used as the synthesis method to summarise the outcome measures. Meta-analysis was not used in this case due to the anticipated heterogeneity.

## 3. RESULTS

### 3.1 Study Selection

The search strategy identified 76 records, of which 11 were duplicates. Sixty-five were screened by reviewing the titles and abstracts and 51 were excluded, and 8 were retrieved for full text review. Three studies met the inclusion criteria (see Table 2). The selection process is summarised in Figure 1. Cohen’s kappa statistics measuring interrater agreement for titles and abstracts, and full-text articles were 1 and 1, respectively.

**Figure 1:**
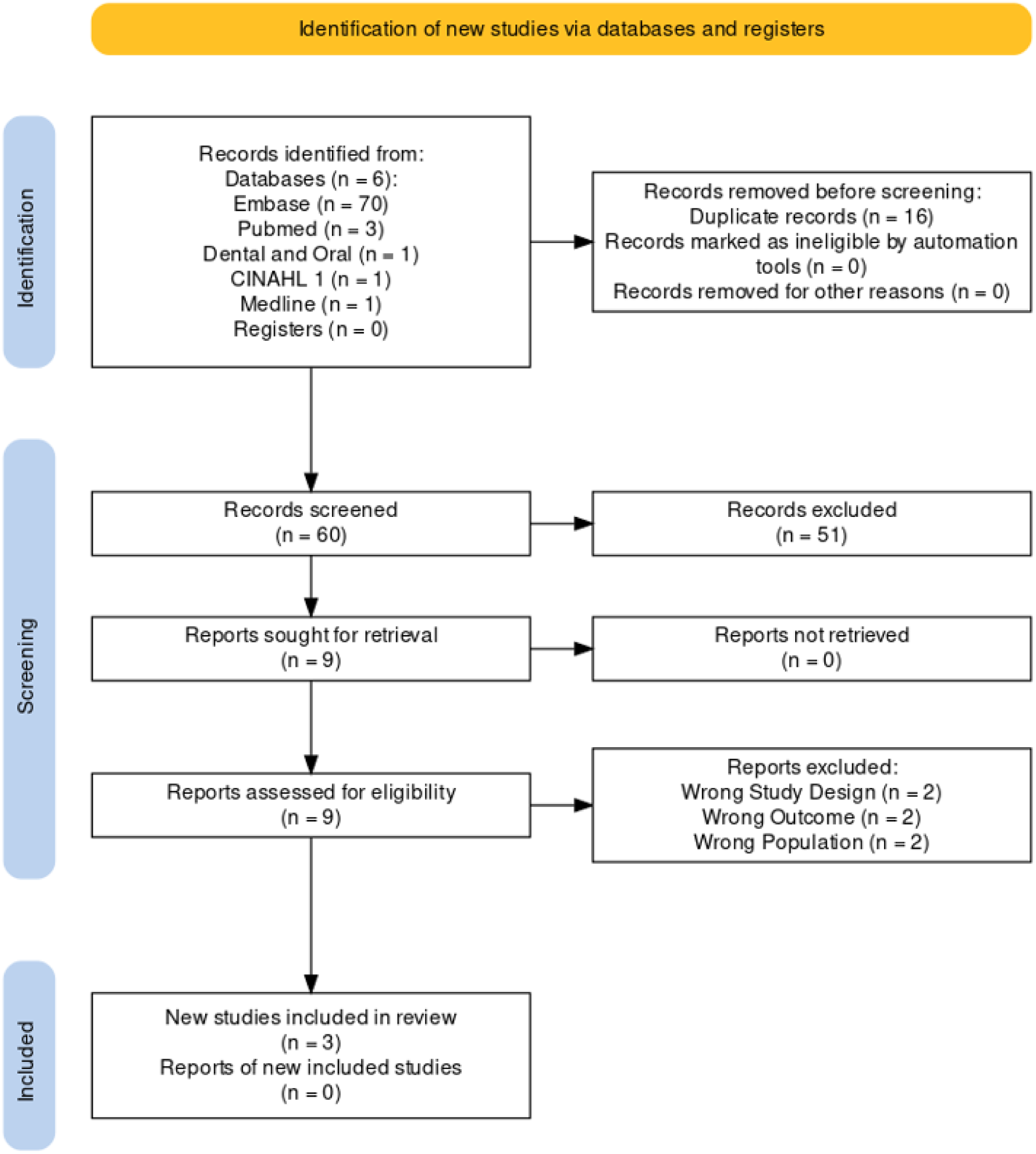
PRISMA Flow Chart.

### 3.2 Study Characteristics

Included in this review are two blinded randomised controlled trials^16,17^, and one randomised controlled trial.^18^ The population group consisted of males and females who were diabetic and had moderate to severe periodontitis. The included studies involved a total of 157 participants (range 40-75 per study), of which 98 participants were randomly assigned to an intervention group and 59 were assigned to the control group. The typical characteristics of the studies are shown in Table 3.

**Table 3:**
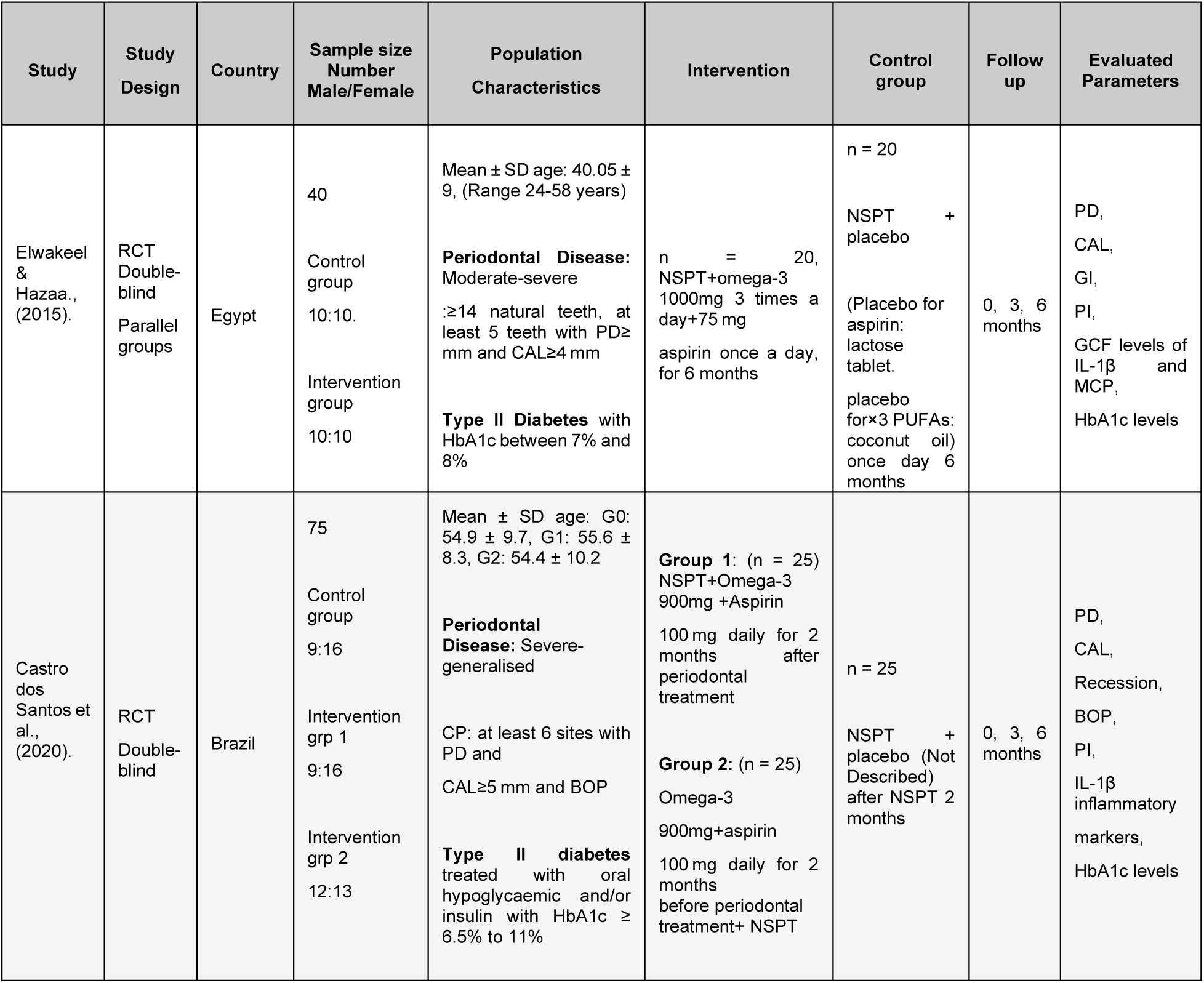
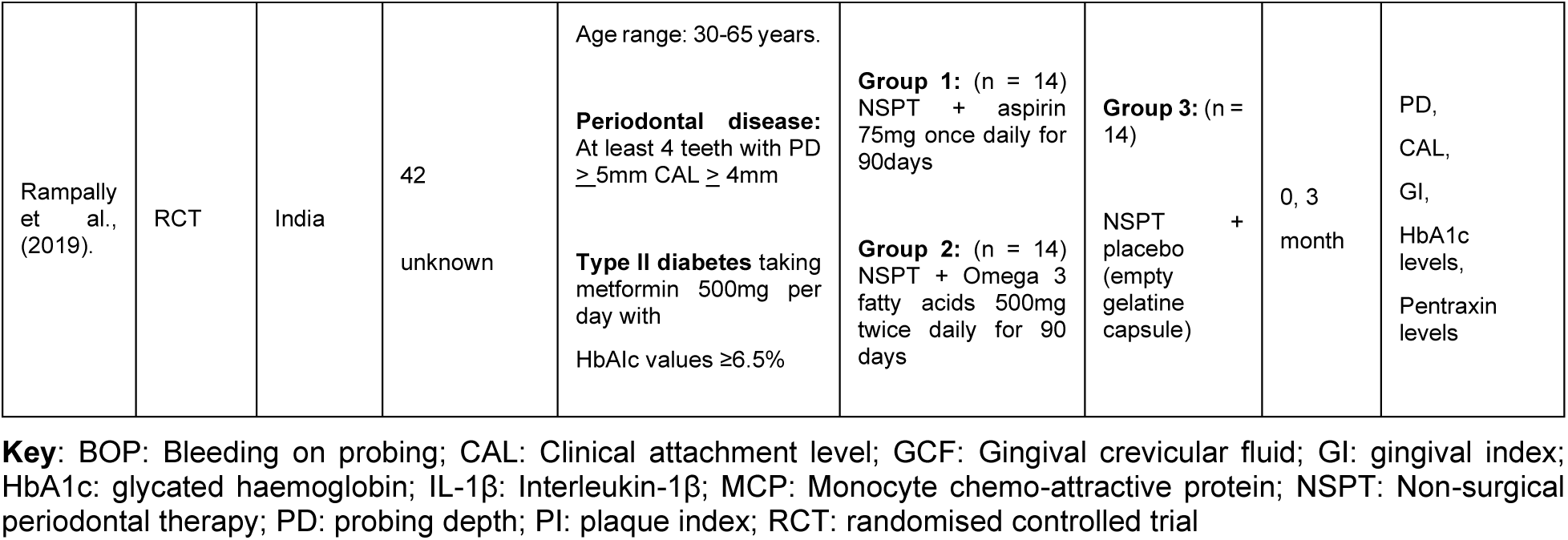
Summary of papers.

### 3.3 Omega 3 and aspirin

Two studies investigated the effect of omega-3 and aspirin as an adjunct to NSPT.^16,17^ Whereas the third investigated these interventions administered separately.^18^ The control group in all three papers were given placebo tablets alongside NSPT for the specified time frames. Only two papers specified the contents of the placebo.^16,18^ The dosage of aspirin ranged between 75 - 100 mg daily and omega-3 ranged between 500 - 3000 mg daily in all. The duration of the administered intervention ranged between 2-6 months.

### 3.4 Non-Surgical Periodontal therapy

Elwakeel et al.^16^ performed NSPT with hand scalers and ultrasonic instrumentation in two sessions, over a two-week period. Whereas Castro dos Santos et al.^17^ provided this treatment over one session. Both studies reported NSPT was completed by a single periodontist who was blinded to the allocation groups at baseline, and each follow-up appointment. Oral hygiene instruction was given and reinforced at every visit. However, Rampally et al.^18^ did not report on how NSPT was performed and whether oral hygiene instruction was given.

### 3.5 Clinical Parameters

Two studies^16,18^ used a Williams probe whereas the third study^17^ used the UNC15 probe to assess the periodontal condition. In each study, PD and CAL were recorded at baseline and at each follow up. Two studies^16,17^ reported that clinical indices were recorded by the same examiner, who remained blinded to allocation and had partaken in calibration exercises (^17^: 92%; ^16^: k value 0.82). The test group in the study by Elwakeel et al.^16^ showed a significant decrease in PD and CAL compared to the control group (*p* < 0.01). Two papers^17,18^ showed a significant improvement between all groups with regards to clinical parameters but failed to establish a statistically significant difference between control and test group (see Table 4). Bleeding on probing (BOP), plaque and gingival indices were not collated for this systematic review due to insufficient data to compare for each study.

**Table 4:**
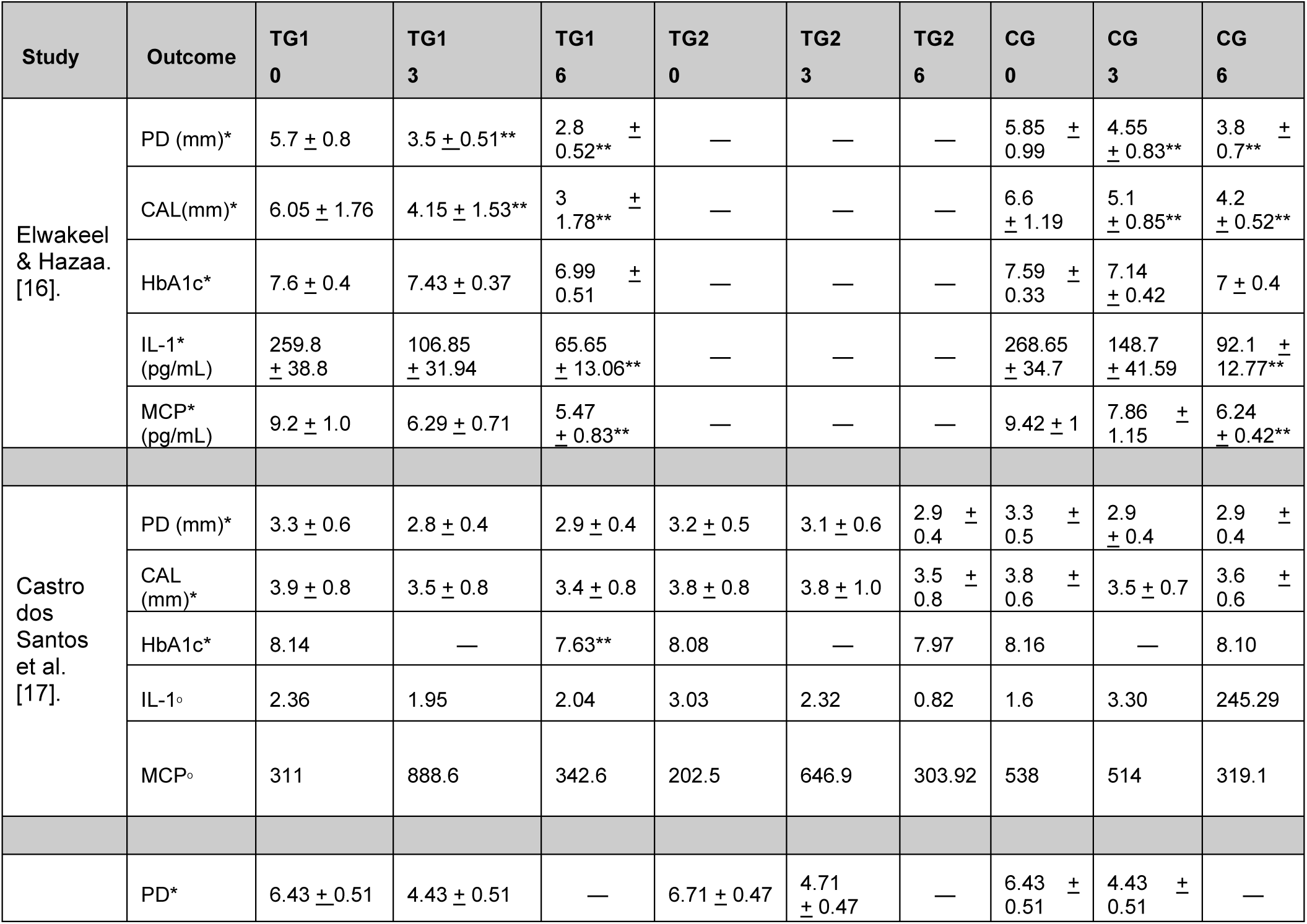
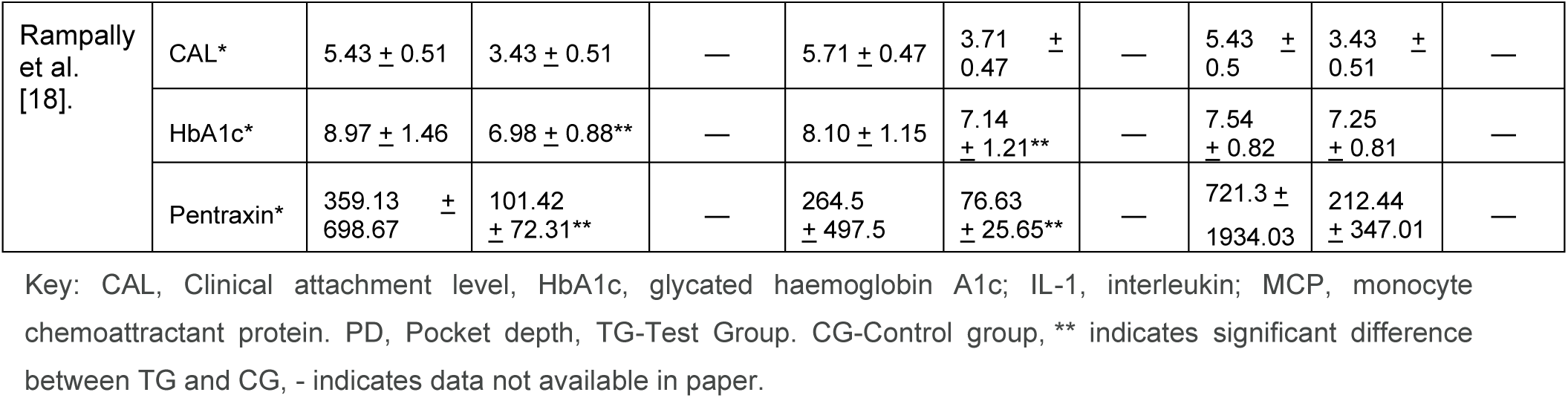
Overview of results.

### 3.6 Biochemical effects

Two studies^16,17^ reported the measurement of multiple inflammatory cytokines in the GCF as IL-1β and MCP were reported in both studies only this data was collated. IL-1β decreased over time in both studies. However, only one study^16^ showed a statistically significant difference compared to the control group (*p* < 0.01). No significant differences were detected in MCP concentration levels in Castro dos Santos et al.^17^, whereas Elwakeel et al.^16^ detected a statistically significant difference (*p* < 0.01) in its MCP levels at 3 and 6 months compared to baseline. Rampally et al.^18^ differed due to the absence of measuring inflammatory cytokines, instead measuring plasma pentraxin (PTX3) which belongs to the C-reactive protein. The greatest improvement was observed in Test group 2, followed by Test group 1 and both indicative of a statistically significant difference (*p* < 0.001) compared to the CG (see Table 4).

### 3.7 Metabolic Parameters

HbA1c were measured at various time points as shown in table 4 for each study. There was a marked improvement in HbA1c values across all three studies; however, only one study^17^ demonstrated a statistically significant difference (*p* < 0.05) compared to the CG.

### 3.8 Risk and quality assessment of bias

Of the three studies included in this systematic review, two scored a low risk of bias and one scored some concerns (see Table 5). Rampally et al.^18^ did not report on the randomisation process, allocation concealment, or blinding. Furthermore, the included studies only reported P-values and did not include confidence intervals or effect sizes.

**Table 5:**
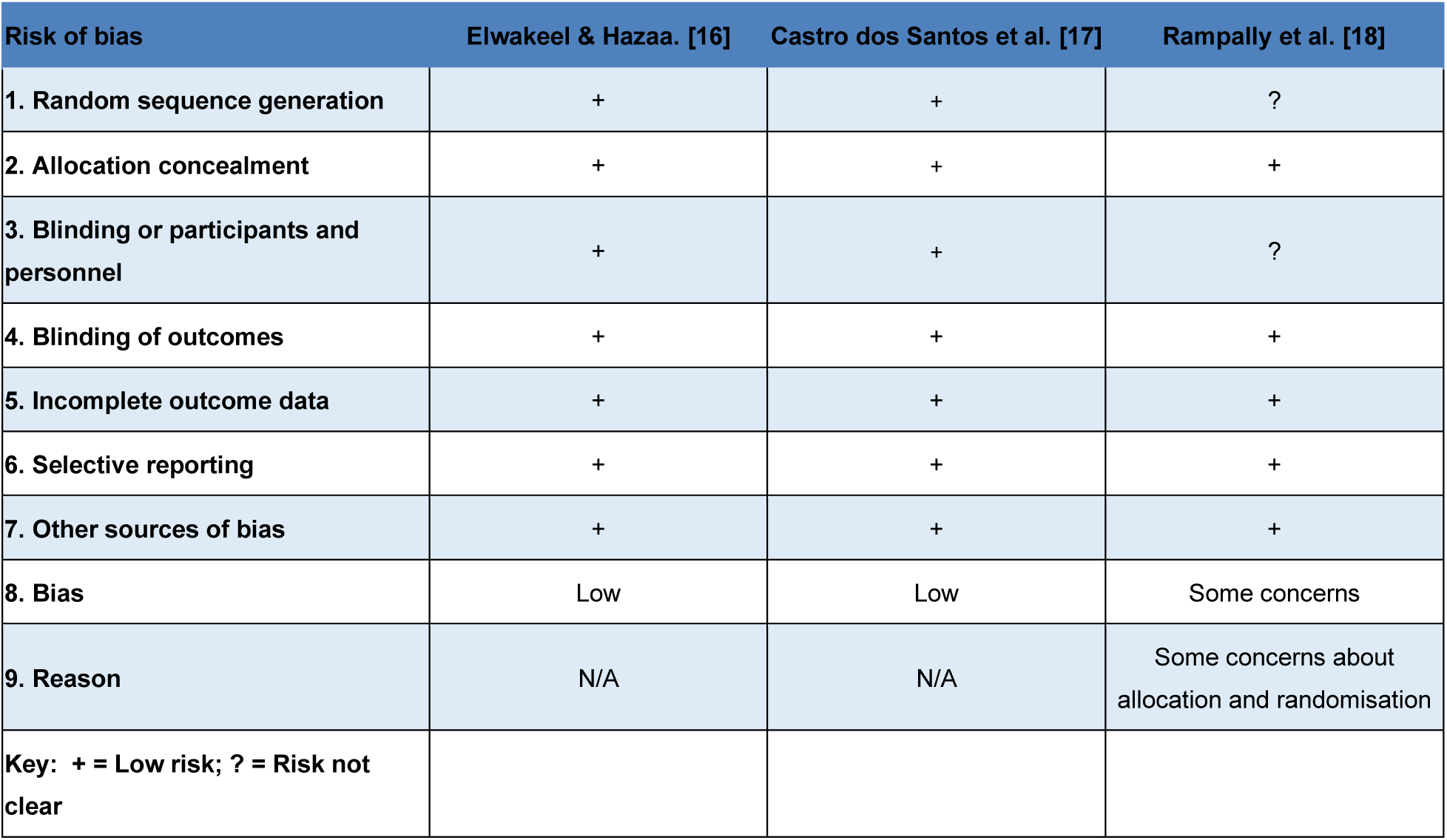
Risk of bias assessment of randomised clinical trials.

## 4. DISCUSSION

### 4.1 Summary of the evidence

This systematic review aimed to investigate the effect of omega-3 and aspirin as adjuncts to non-surgical periodontal therapy (NSPT) in diabetic type II patients with periodontal disease. The review yielded three main findings. Firstly, Elwakeel et al.^16^ reported a statistically significant difference in periodontal clinical parameters and inflammatory markers (*p* < 0.01) when NSPT was performed alongside adjunctive omega-3 and aspirin compared to the control group. Secondly, Rampally et al.^18^ measured pentraxin levels and demonstrated a statistically significant difference compared to a control group (*p* < 0.001). Finally, two studies^17,18^ showed a significant improvement in glycated haemoglobin levels (*p* < 0.05) compared to the control group.

Clinical benefits of adjunct usage of omega-3 and aspirin have been observed in previous studies. El-Sharkawy et al.^19^ proposed omega-3 and aspirin given for 6 months adjunct to NSPT, documented a greater reduction in PD and CAL in the experimental group compared to a control group. Similarly, Elwakeel et al.^16^ observed statistically significant improvements (*p* < 0.01) in PD and CAL favouring the experimental group after 6 months. In contrast, the remaining studies, despite showing an improvement in these parameters, showed no significant difference between experimental and control groups. An explanation could be due to the dosage of intervention given and the period of time within which it was administered. In the included studies, the dosage of aspirin given daily ranged between 75-100mg, whereas omega-3 varied greatly from 500-3000mg daily. The time frame for the administered interventions in the three studies ranged from 2-6 months. This may explain why two of the papers did not yield a significant difference in clinical parameters.

The concentration levels of pro-inflammatory cytokine IL-1β were measured in two of the included papers.^16,17^ These demonstrated an improvement between all groups for IL-1β; however, only Elwakeel et al.^16^ showed a statistically significant reduction (*p* < 0.01) compared to the control group. This supports the findings by Stadler et al.^20^ whereby NSPT alone was effective in decreasing GCF cytokine levels in periodontitis after 6 months. Alexander et al.^21^ supported the use of omega-3 and aspirin after periodontal flap debridement and demonstrated a significant reduction in levels of IL-1β after 6 months. Only one study measured pentraxin levels (PTX3) which belongs to the family of C-reactive proteins (CRP).^18^ There was a significant difference (*p* < 0.01) in PTX3 levels in both intervention groups compared to the control group. However, the improvement significantly favoured the omega-3 group compared to the aspirin test group when combined with NSPT. CRP serves as an activator for the complement system which assists the innate immune system in the destruction of the periodontium. Telgi et al. ^22^ concluded that NSPT was effective in reducing CRP in GCF; however, this study was conducted on healthy individuals without systemic disease. A more recent systematic review showed dietary omega-3 is associated with lower inflammatory biomarkers in the GCF among diabetics and is a useful therapy for other chronic inflammatory diseases such as cardiovascular disease and rheumatoid arthritis.^23^ Therefore, this highlights the potential benefit of administering host modulation therapy in patients with risk factors such as systemic diseases like diabetes.

NSPT is an effective form of removing pathogenic bacteria from the periodontium, which promotes the healing process. However, the healing process in Type II Diabetes Mellitus is more complicated as it is dependent on the level of glycemic control. Research has shown with each 1% reduction in HbA1c levels the risk for Diabetes Mellitus Complications decrease.^24^ Numerous studies have investigated NSPT on glycaemic control, but the evidence is often conflicting and inconsistent. Some evidence shows positive effects on glycemic control, whereas other researchers have reported NSPT has no effect on glycemic control, besides the improvement on the periodontium.^25^ Three studies in this review demonstrated an improvement in HbA1c levels across all groups. Two papers^17,18^ showed a statistically significant difference (*p* < 0.05) favouring the experimental group compared to control. A review by Engebretson et al.^26^ observed a modest reduction in HbA1c levels; however, it concluded that there was limited confidence due to small sample sizes. Other studies have suggested the reduction in HbA1c after NSPT is due to a decrease in the inflammatory burden of pro-inflammatory cytokines such as IL-1β, IL-6 and TNF-a.^27^ This finding is consistent with an earlier study by Christgau et al.^28^ which observed that non-surgical periodontal therapy (NSPT) facilitated metabolic control and improved insulin sensitivity by reducing levels of inflammatory cytokines. This raises the issue for further research into the potential benefit of using host modulation therapy as an adjunct to NSPT for diabetics in improving glycaemic control.

### 4.2 Quality of the evidence

In this review, two RCTs were considered to have low risk of bias according to the revised Cochrane risk of bias tool for randomised trials (RoB 2). Whereas the third RCT scored some concerns due to selective reporting of how randomisation and blinding were performed. Randomisation methods reported in this review varied and included a simple coin toss and block randomisation. The CONSORT group for reporting clinical trials^29^, however, does not endorse coin tossing as a method of randomisation in an RCT, as it can introduce potential bias. All three RCTs had a control group with a placebo intervention, but only two of these studies^16,18^ gave information on the contents of the placebo used. This could introduce another potential limitation, as the composition of a placebo should be identified in order to determine its influence on the outcome.^30^ In the three studies reviewed, the NSPT approach varied among the participants in terms of frequency, and it remains unclear whether these differences in treatment approach influenced the clinical outcomes observed.

The studies included in this review acknowledged certain limitations in their own research. These limitations highlighted the need for larger sample sizes and well-designed multi-centre studies to provide more robust evidence. One study reported the true benefit of omega-3 and aspirin as an adjunct to NSPT may have surfaced if the intervention had been given for a longer duration. It is evident that the anti-inflammatory effects of this intervention are dose and time dependent. Higher doses of omega-3 and aspirin given for a longer duration have shown to exhibit greater efficacy^23^, which is proven by one study included in this review which saw a reduction in the measured parameters. However, there are certain drawbacks associated with the use of aspirin and omega-3. Prolonged usage of NSAIDs and high doses of omega-3 have the potential for haemorrhage and gastro-intestinal upset.^31^

### 4.3 Potential limitations in the review process

This review aimed to only investigate the effect of omega-3 and aspirin on diabetic patients. Nonetheless, the comprehensive search strategy uncovered three papers allowing full confidence that all the relevant research was included in this review. English language papers were specified in the inclusion criteria, which could have introduced language bias in this review.

### 4.4 Implications for practice

Current guidelines in the United Kingdom state that NSAIDs and omega-3 given individually as an adjunct to NSPT are not recommended. However, Sanz et al.^32^ suggests that a combination of omega-3 and low-dose aspirin warrants further assessment in managing periodontitis. This viewpoint aligns with the opinions of other researchers such as Geisinger et al.^33^ and Kruse et al.^30^ who emphasise the need for future studies to investigate the use of adjunctive aspirin therapy and omega-3, either individually or in combination, as adjuncts to NSPT in periodontitis patients. These studies highlight the potential value of exploring these interventions further to enhance periodontal treatment outcomes.

### 4.5 Implications for future research

Despite NSPT being the focus for successful treatment outcomes in periodontitis, adjunct HMT could be indicated in the presence of systemic disease such as diabetes, where NSPT alone may not be enough to decrease clinical, biochemical, and metabolic parameters. Future studies in primary care may be more favourable and should consider or address the methodological limitations of the current published literature. Other recommendations for future research include well-designed RCTs with more uniformity assessing the dosage and duration of the intervention. Evidence reported in clinical trials would be improved by following the CONSORT guidelines.

## 5. CONCLUSION

Omega-3 and aspirin have the potential to decrease clinical parameters and inflammatory markers in the GCF. When administered as an adjunct to NSPT may also improve glycemic control and contribute to the management of type II diabetes. However, it is evident that there were conflicting results as not every paper demonstrated a statistically significant difference in each outcome. As discussed, possible reasons for these results may include the intervention being dose-duration dependent, small sample sizes, and limited evidence currently available. Therefore, due to the uncertainty and limited availability of clinical data at present, there is insufficient evidence to support the use of omega-3 and aspirin as adjuncts to periodontitis in Type II Diabetes Mellitus. Further research addressing the limitations of the current literature is needed.

### 5.1 Implications for Clinical Practice

The potential benefits of omega-3 and aspirin as adjuncts to NSPT include reductions in periodontal clinical parameters and inflammatory markers, improving periodontal health and possibly glycaemic control. However, inconsistent results and limited evidence mean that their integration into clinical practice should be approached cautiously and requires further validation.

### 5.2 Implications for Research

Future research should focus on addressing current limitations, including small sample sizes, varying dosages and durations, and inconsistent reporting. Well-designed randomised controlled trials with greater uniformity are necessary to clarify the efficacy of these adjuncts and provide stronger clinical recommendations.

## Data Availability

All data produced in the present work are contained in the manuscript

## ACKNOWLEDGMENTS

No additional acknowledgments beyond those of the authors have been made.

## Funding and Ethics

No additional acknowledgments beyond those of the authors have been made, and no funding was provided for this research. No ethical approvals were required for this systematic review.

